# GLUCOCOVID: A controlled trial of methylprednisolone in adults hospitalized with COVID-19 pneumonia

**DOI:** 10.1101/2020.06.17.20133579

**Authors:** Luis Corral-Gudino, Alberto Bahamonde, Francisco Arnaiz-Revillas, Julia Gómez-Barquero, Jesica Abadía-Otero, Carmen García-Ibarbia, Víctor Mora, Ana Cerezo-Hernández, José L. Hernández, Graciela López-Muñíz, Fernando Hernández-Blanco, Jose M. Cifrián, Jose M. Olmos, Miguel Carrascosa, Luis Nieto, María Carmen Fariñas, José A. Riancho, for the GLUCOCOVID investigators, Alberto Bahamonde, Fernando Hernández-Blanco, Cristina Buelta-González, Luis A. Marcos-Martínez, Ana I. Martínez-Vidal, Pilar R.l Dosantos-Gallego, Jesús Pérez-Sagredo, Silvia Sandomingo-Freire, Rebeca Muñumer-Blázquez, Antonio Paredes-Mogollo, Elena Brague-Allegue, Miguel Carrascosa, Juan L. García-Rivero, José A. Riancho, José M. Olmos, Carmen Fariñas, José M. Cifrian, Carmen García-Ibarbia, Jose L. Hernández, Francisco Arnaiz-Revillas, Victor Mora, Sara Nieto, Juan Ruiz-Cubillán, Arancha Bermúdez, Javier Pardo, Carlos Amado, Andrés Insunza, Aritz Gil, Teresa Diaz-Terán, Marina Fayos, Miguel A. Zabaleta, Juan J. Parra, Luis Corral-Gudino, Julia Gómez-Barquero, Jesica Abadía-Otero, Ana Cerezo-Hernández, Graciela López-Muñíz, Angela Ruíz-de-Temiño-de-la-Peña, C. Ainhoa Arroyo-Domingo, Javier Mena-Martín, Pablo Miramontes-González, Ana E Jiménez-Masa, Luis Pastor-Mancisidor, Tanía M Álvaro-de-Castro, María Cruz Pérez-Panizo, Tomás Ruíz-Albi, C Gema de-la-Colina-Rojo, María Andrés-Calvo, Andrea Crespo-Sedano, Begoña Morejón-Huerta, Laisa S. Briongos-Figuero, Julio F Frutos-Arriba, Javier Pagán-Buzo, Miriam Gabella-Martín, Marta Cobos-Siles, Ana Gómez-García, Luis Nieto

## Abstract

**Background:** We aimed to determine whether a 6-day course of intravenous methylprednisolone (MP) improves outcome in patients with SARS CoV-2 infection at risk of developing Acute Respiratory Distress Syndrome (ARDS).

**Methods:** Multicentric, partially randomized, preference, open-label trial, including adults with COVID-19 pneumonia, impaired gas exchange and biochemical evidence of hyper-inflammation. Patients were assigned to standard of care (SOC), or SOC plus intravenous MP [40mg/12h 3 days, then 20mg/12h 3 days]. The primary endpoint was a composite of death, admission to the intensive care unit (ICU) or requirement of non-invasive ventilation (NIV).

**Results:** We analyzed 85 patients (34, randomized to MP; 22, assigned to MP by clinician’s preference; 29, control group). Patients’ age (mean 68±12 yr) was related to outcome. The use of MP was associated with a reduced risk of the composite endpoint in the intention-to-treat, age-stratified analysis (combined risk ratio -RR-0.55 [95% CI 0.33-0.91]; p=0.024). In the per-protocol analysis, RR was 0.11 (0.01-0.83) in patients aged 72 yr or less, 0.61 (0.32-1.17) in those over 72 yr, and 0.37 (0.19-0.74, p=0.0037) in the whole group after age-adjustment by stratification. The decrease in C-reactive protein levels was more pronounced in the MP group (p=0.0003). Hyperglycemia was more frequent in the MP group.

**Conclusions:** A short course of MP had a beneficial effect on the clinical outcome of severe COVID-19 pneumonia, decreasing the risk of the composite end point of admission to ICU, NIV or death.

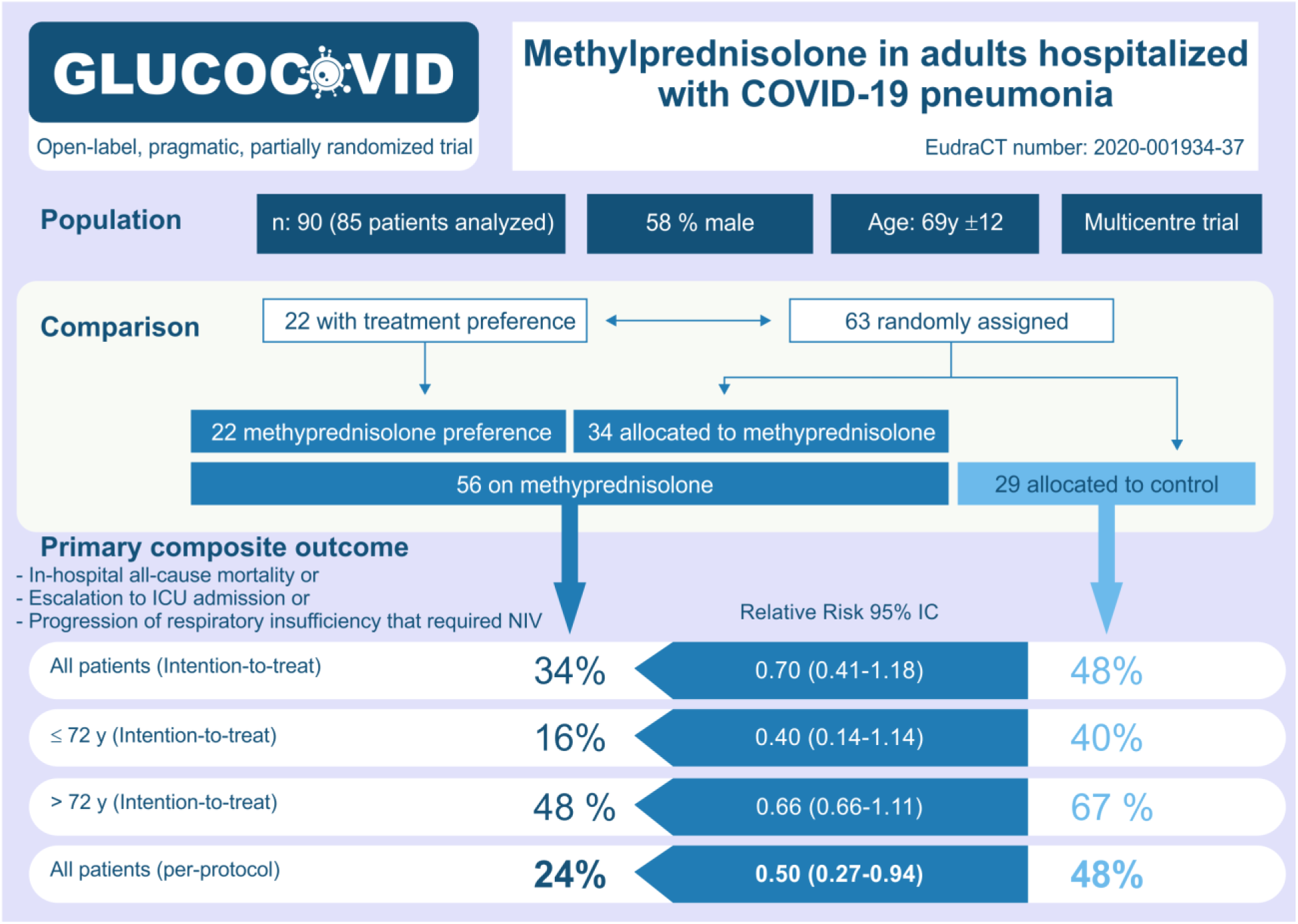

## INTRODUCTION

Since December 2019 the world faces a pandemic, coronavirus disease 2019 (COVID-19), caused by the novel Severe Acute Respiratory Syndrome Coronavirus 2 (ARS-CoV-2). The rapid spread and magnitude of COVID-19, along with the severity of the disease in some patients have stressed the whole world and have put into question our conceptions about viral respiratory infections.

The spectrum of COVID-19 ranges from asymptomatic patients or mild disease to severe progressive pneumonia, with multiple organ failure and death [1]. Patients with severe COVID-19 develop, usually after a first stage with mild manifestations, a disorder similar to acute respiratory distress syndrome (ARDS). These patients suffer a hyper-inflammatory syndrome characterised by a rapid hypercytokinemia targeting the lung parenchyma and/or vasculature [2][3]. This cytokine storm-like state is characterised by increased interleukins (IL) and acute phase reactants [4]. Recent retrospective studies confirmed the association of elevated ferritin, lactate dehydrogenase or IL-6 with poor prognosis [5], thus suggesting that mortality may be related to virally-driven hyper-inflammation. Hence, in this phase, the use of immunomodulators may be justified.

From previous coronavirus outbreaks, such as Severe Acute Respiratory Syndrome (SARS) and Middle East Respiratory Syndrome (MERS), as well as from other viral pneumonias, we learned that corticosteroid therapy should not be routinely recommended, for they might exacerbate lung injury and even increase mortality [6]. Thus, current interim guidance from the World Health Organization (WHO) advises against corticosteroid use in Covid-19 patients unless indicated for another reason [7].

However, the rapid progression of severe cases of SARS-CoV-2 infection, along with the marked increase in several laboratory biomarkers of systemic inflammatory response and the absence of effective antiviral therapy, has led clinicians to question the recommendation against using corticosteroids. Besides, the potential benefit of corticosteroids in ARDS of other causes prompted interest in using them in COVID-19 patients [8]. Thus, corticosteroids and other immunomodulators are now frequently used in severe COVID-19 cases [9,10] and have gained support from some scientific societies under certain circumstances [11,12]. Recommendations to prescribe corticosteroids are based on anecdotal observations and retrospective uncontrolled series of patients, but so far no controlled prospective trials are available [13–16]. For instance, a multicentre study comparing two periods of COVID-19 attendance with or without steroids showed a beneficial effect of the early use of corticosteroids [16]. Interestingly, a composite end-point (escalation of care from the hospitalization ward to the intensive care unit (ICU), new requirement for mechanical ventilation, or mortality) occurred in 54% of patients who received standard therapy, and in 35% of those treated with corticosteroids (p=0.005). In a retrospective study of 201 patients, methylprednisolone (MP) was associated with reduced mortality in patients with ARDS [14]. In another retrospective study, 11% of patients on MP and 35% patients without corticosteroids required mechanical ventilation (p=0.05) [17]. On the contrary, some studies argued that corticosteroids may be deleterious and cause a delayed viral clearance in COVID-19 [18], as it was also found in SARS [19].

Up to 5-10% of the hospitalized patients with COVID-19 develop ARDS and require respiratory support in ICU [20]. Lacking a drug specifically designed for this novel coronavirus and with the prospect of several months or even years until the development of an effective vaccine, we urgently need some drug repositioning for the treatment of COVID-19. These considerations motivated us to design and conduct a pragmatic, randomized, controlled trial (GLUCOCOVID) to explore the role of a short course of MP in patients with COVID-19 pneumonia at risk of developing respiratory failure and ARDS. Here we report a planned interim analysis of the first 90 patients included.

## METHODS

### Study design

GLUCOCOVID is a partially randomized preference, open-label, controlled, two-arm, parallel-group, trial conducted at 5 hospitals in Spain in April-May 2020. The study was designed to address the efficacy of adding corticosteroids to standard therapy in patients with moderate-severe COVID-19.

We designed a pragmatic, partially randomized trial, including a clinician’s preference arm in an attempt to avoid inclusion bias in the current setting in which many physicians feel glucocorticoids may have a beneficial effect in COVID-19 despite the absence of controlled clinical trials. This approach is based upon the well-described preference trial designs, which allow incorporating individual’s preferences and questions about equipoise [21,22]. The principal investigator of every hospital encouraged the medical team to maximize the number of patients included by the randomization way, but all included patients were analysed regardless they were randomized or not [23]. The study was registered at the European Clinical Trials Register (EudraCT number: 2020-001934-37) and the Spanish Registry of Clinical Studies (2020-001934-37).

### Participants

Eligible patients were hospitalized subjects over 18 years of age, with a laboratory confirmed diagnosis of SARS-CoV2 infection. Additional inclusion criteria were all the following:

1. Symptom duration of at least 7 days
2. Radiological evidence of lung disease in chest X-ray or CT-scan
3. Moderate-to-severe disease with abnormal gas exchange: PaFi (PaO2/FiO2) < 300, or SAFI (SAO2/FiO2) < 400, or at least 2 criteria of the BRESCIA-COVID Respiratory Severity Scale (BCRSS) [24].
4. Laboratory parameters suggesting a hyper-inflammatory state: serum C-Reactive Protein (CRP) >15 mg/dl, D-dimer > 800 mg/dl, ferritin > 1000 mg/dl or IL-6 levels > 20 pg/ml.

Patients were excluded if they were intubated or mechanically ventilated, were hospitalized in the ICU, were treated with corticosteroids or immunosuppressive drugs at the time of enrollment, have chronic kidney disease on dialysis, were pregnant or refused to participate.

The study was approved by the Institutional Review Boards of participating hospitals, and patients gave informed consent.

### Treatment allocation

Once an eligible patient was identified, if the clinical team decided that a strong preference for glucocorticoid therapy existed, the patient was allocated to the preference arm. Otherwise, the patient was randomized (1:1) and allocated to the MP or control arm accordingly. Patients were randomized based on a spreadsheet that transformed every medical record number into a group allocation.

### Interventions

Patients in both study groups received standard of care (SOC) therapy according to the local protocols. SOC protocols were similar across the participating hospitals and were based on the Spanish Ministry of Health, Consumer Affairs and Social Welfare technical documents [25] and WHO recommendations [7]. SOC included symptomatic treatment with acetaminophen, oxygen therapy, thrombosis prophylaxis with low molecular weight heparin, and antibiotics for co-infections. Azythromycin, hydroxychloroquine and lopinavir plus ritonavir were frequently prescribed.

Biochemical tests and image studies were performed according to clinical criteria and local protocols, using standard techniques.

In addition to SOC, patients in the experimental group received methylprednisolone (MP) 40 mg intravenously every 12 hours for 3 days and then 20 mg every 12 hours for 3 days. The clinical teams freely prescribed Interleukin-blocking agents and other therapies, as indicated.

### Outcome

The primary outcome measure was a composite endpoint that included in-hospital all-cause mortality, escalation to ICU admission, or progression of respiratory insufficiency that required non-invasive ventilation (NIV).

The secondary outcomes were the effects on the individual components of the composite endpoint and laboratory biomarkers at baseline and 6 days after inclusion (time window 4-8 days).

### Sample size and Statistical analysis

The initial sample size target was estimated assuming that MP could reduce the primary composite endpoint by 50 % or more. With an event rate of 40% in the control arm, 90 patients in each study arm would be needed. Here we report the results of the interim analysis, which was planned a priori after inclusion of one-half of the patients, to avoid delaying the communication of clinically useful data in the current pandemic scenario.

Continuous variables were compared using Student t-test and ANOVA, or Mann-Whitney U test and Kruskal-Wallis test if not normally distributed. We compared categorical variables using Fisher’s exact test. Relative risk ratio and differences in absolute risks were derived from the estimated risks of the primary composite endpoint. In stratified analyses, the combined risk ratio was computed with the Mantel-Haenszel method. Multivariate-adjusted risk ratio was estimated by using unconditional logistic regression. Survival plots were built with the Kaplan-Meier method and compared with the log-rank test. Patients were censored at hospital discharge or day 15 after inclusion.

The analyses were performed according to both intention-to-treat and per-protocol principles. For the latter, we considered those patients in the MP group who had received at least 3 doses of the drug (thus elapsing at least 24 hr from inclusion) before the primary endpoint occurred. The treatment arms were studied considering independently the preference and randomization arms, as well as combining both MP arms.

## RESULTS

Five out of 90 patients initially included were later excluded from the analysis (2 were previously on corticosteroids, 1 was on NIV, 1 was taken to ICU simultaneously to MP onset, and 1 patient with initial suspicion of COVID-19 was finally diagnosed of vasculitis). Thus, 85 patients were analyzed; 22 received MP according to the clinician’s preference, and 63 were randomized. Although allowed by design, no patient was included in the control arm by clinician’s preference. In 3 patients of the control group, but none in the MP group, clinicians prescribed MP boluses after initial allocation because of deterioration of the patient’s condition. The baseline characteristics of the patients are shown in table 1. Those in the MP arm were slightly older, but the baseline characteristics were otherwise very similar across groups. Therefore, we combined the preferential and randomized arms of MP for further analysis (figure 1). The use of lopinavir/ritonavir was slightly more frequent in the control arm. More than 90% of the patients took hydroxichloroquine and/or azithromycin during hospital admission. Although the active search for arrhythmias was not planned in the study protocol, no clinically significant arrhythmias were reported.

**Table 1.** Baseline characteristics of the study groups, Mean and SD for continuous variables, and number and percentage for categorical variables.

|  | Total<br>(n=85) | Preference arm<br>(methyl-<br>prednisolone)<br>(n=22) | Control arm<br>(n=29) | Methyl-<br>prednisolone<br>arm<br>(n=34) | p |
| --- | --- | --- | --- | --- | --- |
| Age, yr | 69±12 | 66±13 | 66±12 | 73±11 | 0.03 |
| Sex (male, %) | 49 (58) | 10 (45) | 16 (55) | 23 (68) | >0.10 |
| COVID-19 characteristics |  |  |  |  |  |
| • SAFI (SaO<sub>2</sub>/FI O<sub>2</sub>) | 285±105 | 323±78 | 281±121 | 264±101 | >0.10 |
| • Creatinine, mg/dl | 0.9±0.4 | 1.0±0.4 | 0.9±0.4 | 0.8±0.3 | 0.10 |
| • Lymphocytes/μl | 872±760 | 752±21 | 814±305 | 997±1125* | >0.10 |
| • Platelets /μl | 293764±<br>353413 | 255681±<br>135722 | 269827±<br>96344 | 261205±<br>139107 | >0.10 |
| • CRP, mg/dl | 16.1±8.4 | 12.8±8.4 | 17.2±8.8 | 17.1±7.8 | >0.08 |
| • D Dimer, mg/dl | 2308±5395 | 2310±3638 | 1297±982 | 3163±7902 | >0.10 |
| • Ferritin, mg/dl | 1129±905 | 992±945 | 1078±882 | 1259±907 | >0.10 |
| Comorbidities |  |  |  |  |  |
| • Hypertension, n (%) | 39 (46) | 9 (41) | 12 (41) | 18 (53) | >0.10 |
| • Cardiac disease, n (%) | 9 (11) | 1 (5) | 4 (14) | 4 (12) | >0.10 |
| • Respiratory disease, n (%) | 7 (8) | 2 (9) | 1 (3) | 4 (12) | >0.10 |
| • Diabetes, n (%) | 13 (15) | 2 (9) | 4 (14) | 7 (21) | >0.10 |
| Therapy |  |  |  |  |  |
| • Azithromycin, n (%) | 76 (89) | 18 (81) | 29 (100) | 29 (85) | 0.07 |
| • Hydroxychloroquine, n (%) | 81 (95) | 20 (91) | 29 (100) | 32 (94) | >0.10 |
| • Lopinavir/Ritonavir, n (%) | 67 (79) | 14 (63) | 28 (97) | 25 (74) | 0.01 |

- Included 1 patient with CLL

**Figure 1.**
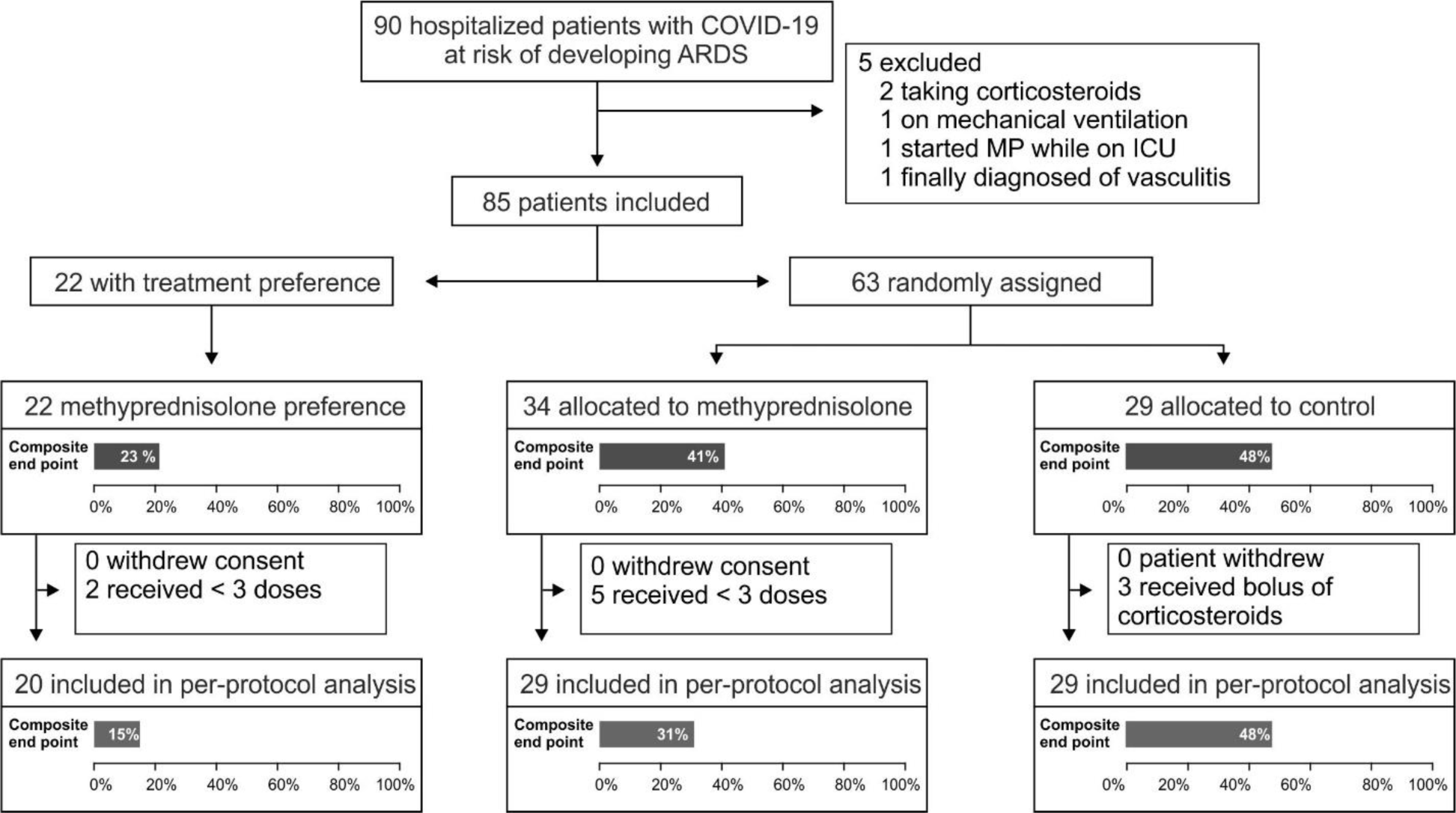
GLUCOCOVID flow diagram

In the intention-to-treat analysis, all patients who received at least one dose of MP were included in the treatment arm. In this univariate analysis, age and baseline SAFI and CRP levels were the only variables significantly associated with the primary composite endpoint (admission to ICU, NIV, or death). Mean age was 67±11 and 72±13 yr in patients with a good and a bad outcome, respectively (p=0.07); mean SAFI was 327±93 and 218±86, respectively (p<0.001); and mean CRP was 14.3±8.3 and 18.8±8.0, respectively (p=0.016). In line with the influence of patients’ age on disease severity, subjects above the median age of 72 yr were more likely to reach the composite endpoint than those aged 72 yr or less (relative risk 1.97, 95% CI 1.11-3.47, p=0.025). The primary composite endpoint occurred somewhat less frequently in the MP group. Although the difference was not statistically significant in the unadjusted analysis, in the age-stratified analysis, MP was associated with a significantly lower risk of bad outcome, with a 45% relative risk reduction and 24% absolute risk reduction (table 2). Thus, age was a confounding variable, but there was no statistically significant interaction between age and the effect of MP.

**Table 2.** Intention-to-treat analysis. Comparison of patients in the control and methylprednisolone (MP) arms. Unstratified and age-stratified analyses.

|  | **Adverse outcome\*** | **Good outcome** | **Relative Risk** | **p** |
| --- | --- | --- | --- | --- |
| **All patients** |  |  |  |  |
| Control | 14 (48) | 15 (52) | 1 |  |
| MP | 19 (34) | 37 (66) | 0.70<br>(0.41-1.18) | 0.25 |
| **Age ≤72** |  |  |  |  |
| Control | 8 (40) | 12 (60) | 1 |  |
| MP | 4 (16) | 21 (84) | 0.40<br>(0.14-1.14) |  |
| **Age >72** |  |  |  |  |
| Control | 6 (67) | 3 (33) | 1 |  |
| MP | 15 (48) | 16 (52) | 0.66<br>(0.40-1.11) |  |
| **Combined (MH)** |  |  | 0.55<br>(0.33-0.91) | 0.025 |

\*Primary composite outcome (ICU admission, NIV or death)
MP: methylprednisolone. MH: Mantel-Haenszel

At baseline, all study groups had similar biomarker levels. Six days later, CRP levels were lower in both groups, but the decrease was more pronounced in the MP group (p=0.0003). Other biomarkers were similar in the control and MP groups (figure 2).

**Figure 2.**
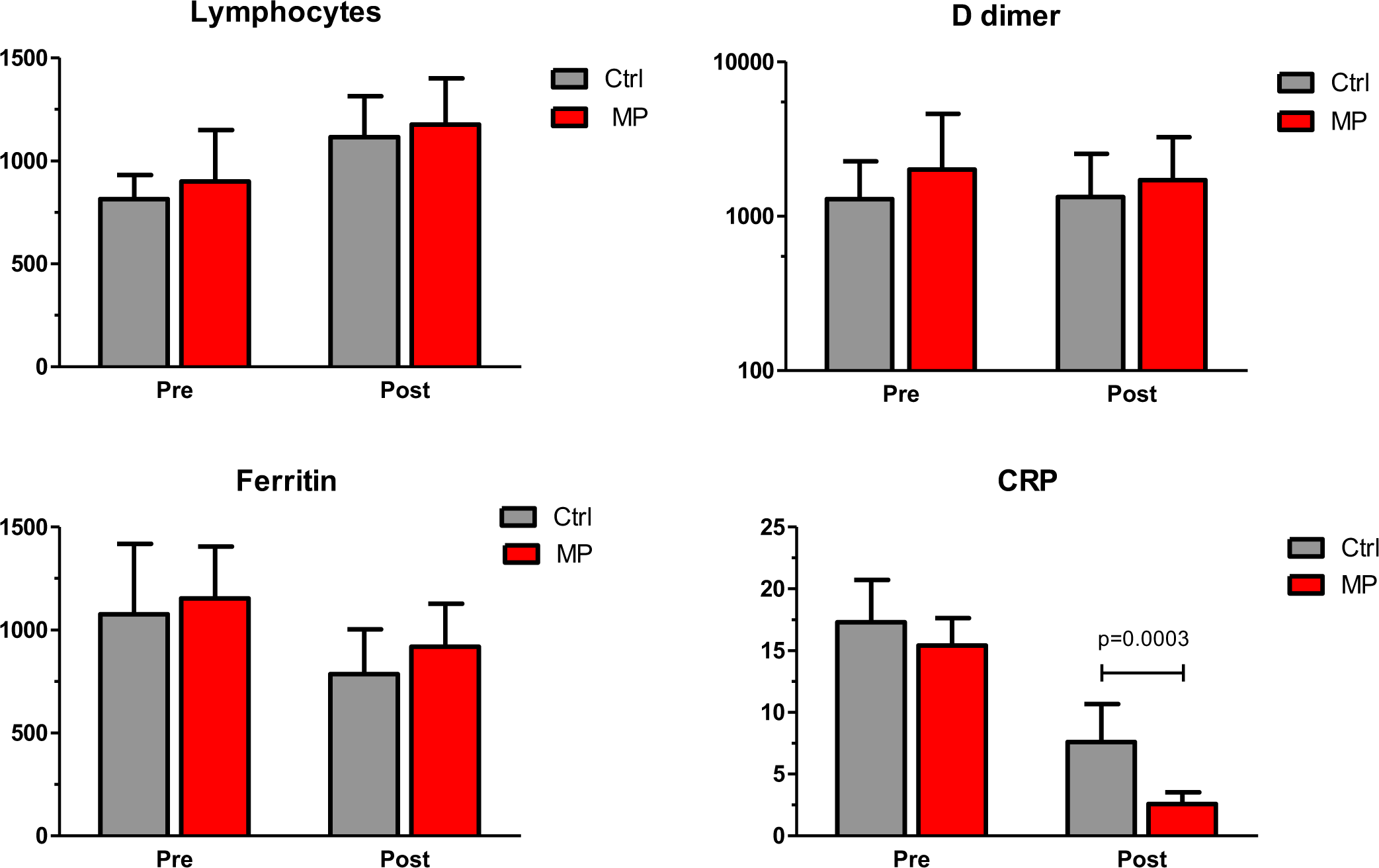
Biomarkers of control and MP groups at baseline (Pre) and 6 days (range 4-8) after inclusion. Mean and 2xSEM.

Fifteen patients received the IL-6 blocking agent tocilizumab (10 [18%] in the MP group and 4 [14%] in the control group); in 7 patients of the MP group (12%), the IL-1 blocking agent anakinra was prescribed. After patients treated with tocilizumab and/or anakinra were excluded from the analysis, the results were similar to those found in the whole group: MP-treated patients had a lower risk of reaching the combined endpoint (Relative risk reduction 55%; 95% CI 15-76%; p=0.015).

No major side effects were observed, but hyperglycemia (>180 mg/dl) was more frequent in the MP group. Twelve patients on MP (21%), and none in the control group developed hyperglycemia >180 mg/dl (p=0.006).

In the per-protocol analysis, we included 78 patients who received at least 3 doses of MP before the composite endpoint occurred (this is, at least 24 hours elapsed between inclusion and the occurrence of an endpoint event). As shown in table 3, MP was associated with a 50% lower risk of an adverse outcome, in the overall analysis, and a 63% relative reduction of adverse outcome risk in the age-stratified analysis. In the multivariate analysis, adjusting by age and baseline SAFI, patients on MP had a relative risk of 0.28 (95% CI 0.08-0.80, p=0.013). Regarding the individual components of the main outcome variable, patients on MP had a significantly lower risk of ICU admission (8% vs 28%, p=0.047), with similar frequency of NIV (6% vs 10%, ns) and death (20% vs 18%, ns) (supplementary table). Similarly, when the analysis was limited to the randomized arms, the risk of adverse outcome, as defined by the composite endpoint, was significantly lower in the MP group than in the control group, with an adjusted risk ratio (adjusted by age and baseline SAFI) of 0.28 (95% CI 0.07-0.90, p=0.029).

**Table 3.**
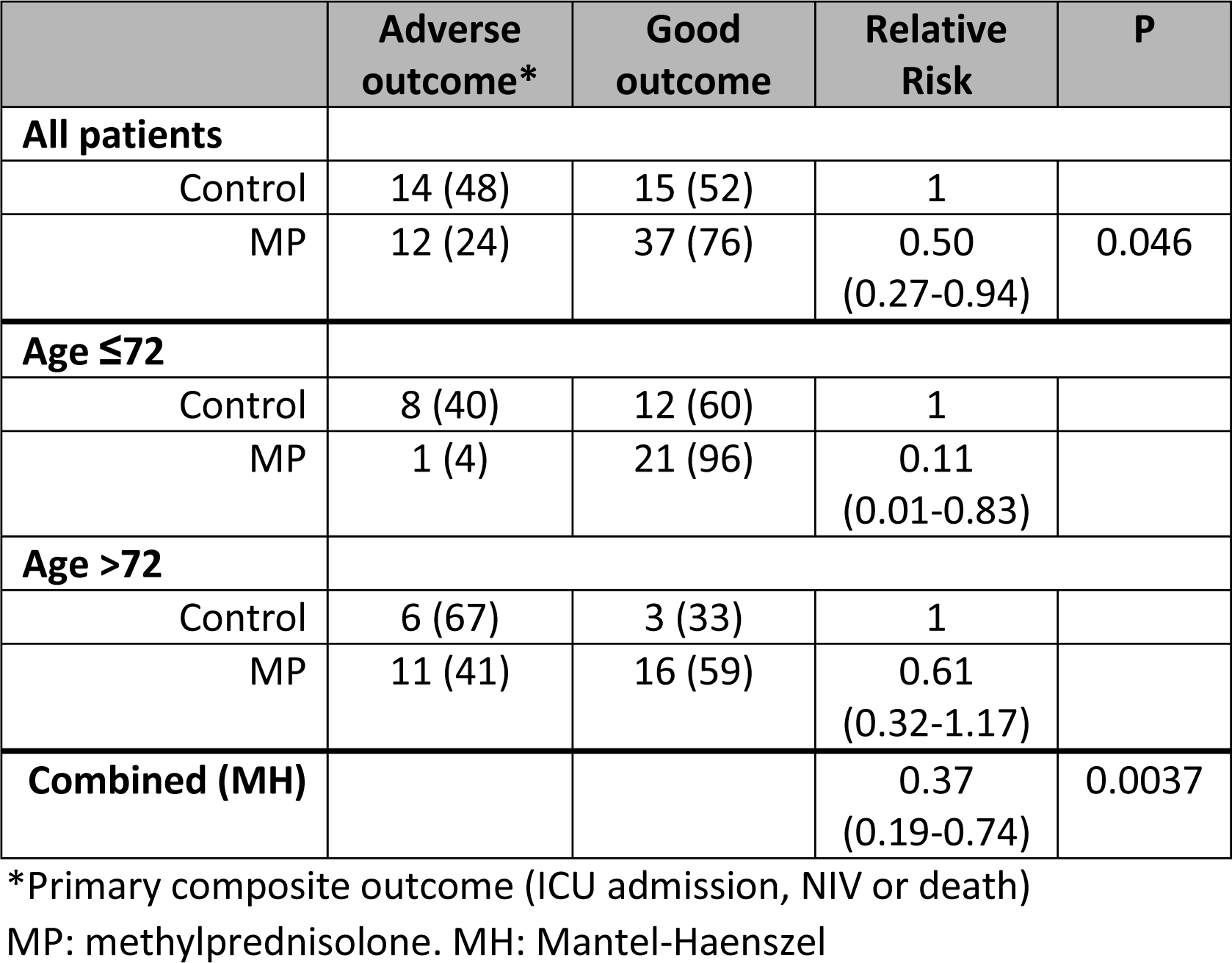
Per protocol analysis after excluding 7 patients who received only 1-2 doses of methylprednisolone.

|  | **Adverse outcome\*** | **Good outcome** | **Relative Risk** | **P** |
| --- | --- | --- | --- | --- |
| **All patients** |  |  |  |  |
| Control | 14 (48) | 15 (52) | 1 |  |
| MP | 12 (24) | 37 (76) | 0.50<br>(0.27-0.94) | 0.046 |
| **Age ≤72** |  |  |  |  |
| Control | 8 (40) | 12 (60) | 1 |  |
| MP | 1 (4) | 21 (96) | 0.11<br>(0.01-0.83) |  |
| **Age >72** |  |  |  |  |
| Control | 6 (67) | 3 (33) | 1 |  |
| MP | 11 (41) | 16 (59) | 0.61<br>(0.32-1.17) |  |
| **Combined (MH)** |  |  | 0.37<br>(0.19-0.74) | 0.0037 |

\*Primary composite outcome (ICU admission, NIV or death)
MP: methylprednisolone. MH: Mantel-Haenszel

Survival analysis confirmed the influence of age and treatment. Patients on MP had a significantly higher chance of good outcome (p=0.001 by log-rank test) (figure 3). Similar results were observed when patients assigned to MP by clinician’s preference and those randomized to MP were considered separately. Pairwise-comparisons revealed significant differences between control and MP groups, but not between both MP groups (control-randomized MP, p=0.019; control-preference MP, p=0.003; randomized MP-preference MP, p=0.233; supplementary figure).

**Figure 3.**
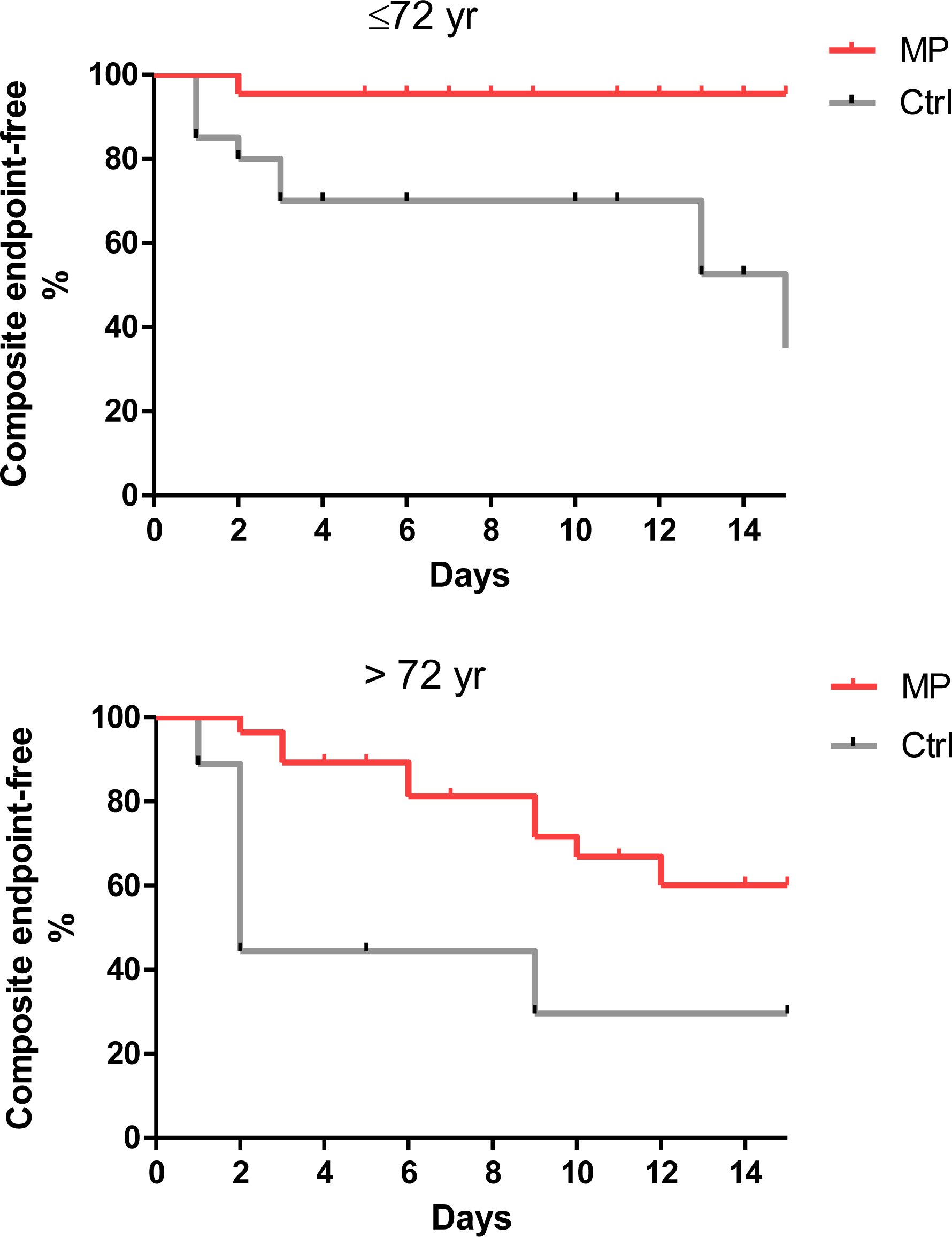
Kaplan-Meier plots showing the probability of not occurring the primary composite endpoint (ICU admission, need of NIV or death) of the control (grey) and MP (red) groups in COVID-19 patients stratified by age.

## DISCUSSION

COVID-19 has put the whole world under unprecedented stress. Thus, clinicians have been forced to take decisions in the absence of solid evidence about diagnosis and therapy. However, an impressive amount of information has been gathered in a few weeks, which has led to deeper disease knowledge and better patient management. For example, from the initial conception of COVID-19 as a pure infectious disease, accumulated data have helped to understand the important role of the host inflammatory response. In this line, uncontrolled observations have suggested a beneficial effect of anti-inflammatory therapy, including IL-blocking agents and glucocorticoids [16,26,27]. The latter are particularly appealing because they are inexpensive, widely available, and easy to administer. However, the role of corticosteroids in viral-induced and other forms of ARDS is controversial [19].

Our study was motivated by this controversy. Since many clinicians in our hospitals had a positive feeling about the effect of glucocorticoids in patients with severe COVID-19, a purely randomized design appeared difficult to follow, and it could likely had implied a high risk of inclusion bias. So, we adopted a pragmatic mixed preference/randomized design. Although this design may complicate the analysis, the three arms showed similar baseline characteristics (except for some differences in patients’ age). In fact, patients in the MP arm were somewhat older than those in the control arm. This is an important issue, for in this study we confirmed that advanced age is a risk factor for poor outcome, in line with previously reported series [14,28–30]. The confounding effect of age is complex, as it may influence not only the course of the disease, but also decisions about escalation to ICU admission in a scenario of limited resources.

Interestingly, in this trial MP administration was associated with a reduced risk of poor outcome, which was statistically significant after adjustment for confounding factors, such as age and baseline respiratory status (as assessed by SAFI). Our results are consistent with those of a recent quasi-experimental study [16] that used similar endpoints and MP doses. The primary composite endpoint occurred in 54% patients in the SOC group and in 35% in the early glucocorticoids group. Those figures are remarkably similar to ours (48% vs 34%).

Our study has several limitations. Firstly, the small sample size. Indeed, it is a pre-planned interim analysis of an ongoing trial, not powered to explore the association of treatment with individual endpoints. However, we feel the results are important to inform clinical decisions while ongoing larger controlled randomized trials are completed. Secondly, the inclusion of a preferential arm theoretically hampers the balance of baseline characteristics across study arms. Nevertheless, actual differences were not large, as shown in table 1. In fact, the beneficial effect of MP was observed not only in the analysis combining the randomized and preferential arms, but also when only the randomized arms were compared, reinforcing the conclusions of the study. Third, there might be differences in patient management across the participating hospitals. Fortunately, in practice, the protocols for COVID-19 were very similar, because they were based on the recommendations of the Spanish Ministry of Health, including the use of azithromycin, hydroxychloroquine, and lopinavir/ritonavir. Fourth, due to the rapidly deteriorating course of some COVID-19 cases, they escalated to ICU or NIV within the first 24 hours of inclusion in the study. Therefore, they received only 1-2 doses of MP, thus impeding to assess the effect of the drug. We included a per-protocol analysis excluding those patients to avoid their confounding effect.

Our study shows that MP improves the prognosis of COVID-19. Elucidating its precise role within a treatment strategy would need further studies, but our data suggest that MP is useful in patients with moderate/severe disease with evidence of inflammatory activation. However, several patients in our cohort deteriorated rapidly and required escalation of therapy. Thus, it would be interesting to initiate studies to explore the role of glucocorticoids at somewhat earlier stages of disease.

In conclusion, the interim analysis of this ongoing clinical trial shows a beneficial effect of a short course of methylprednisolone on the clinical outcome of patients with severe COVID-19. Our data suggest that corticosteroids may have a clinically important effect in reducing the risk of developing severe respiratory insufficiency and ARDS.

## Data Availability

Request from authors

## Acknowledgments

We thank Prof. Jose Luis Pérez-Castrillón (Universidad de Valladolid) for useful discussion and comments on the manuscript, and Dr. Mar García Saiz (Hospital UM Valdecilla) for helping with trial registry.

This trial would have been impossible without the support and collaboration of the hundreds of health professionals involved in the care of COVID-19 patients in our hospitals during the 2020 Spring pandemic.

## Author roles

Conceptualization: LC-G, JLH, MCF, JAR

Data collection: All authors

Data analysis: LC-G, JAR

Manuscript draft: LC-G, MCF, JMO, JMC, FA, VM, AB, JAR

Critical input and final manuscript approval: All authors

Project supervision: LC-G, JAR

JAR has full access to the data and is the guarantor for the data

## Figure legends

**Supplementary table.** Individual components of the composite endpoint. Number and (%). MP: methylprednisolone. ITT: intention-to-treat. PP: per-protocol (>2 doses MP)

|  | Control | MP ITT | MP PP |
| --- | --- | --- | --- |
| All patients | n=29 | n=56 | n=49 |
| Death | 5 (17) | 12 (21) | 9 (18) |
| ICU | 8 (28) | 8 (14) | 4 (8) |
| NIV | 3 (10) | 6 (11) | 3 (6) |
| ≤72 yr | n=20 | n=25 | n=22 |
| Death | 1 (5) | 0 (0) | 0 (0) |
| ICU | 8 (40) | 4 (16) | 1 (4) |
| NIV | 1 (5) | 0 (0) | 0 (0) |
| >72 yr | n=9 | n=31 | n=27 |
| Death | 4 (44) | 12 (42) | 9 (33) |
| ICU | 0 (0) | 4 (13) | 3 (11) |
| NIV | 2 (22) | 6 (19) | 3 (11) |

**Supplementary figure.**
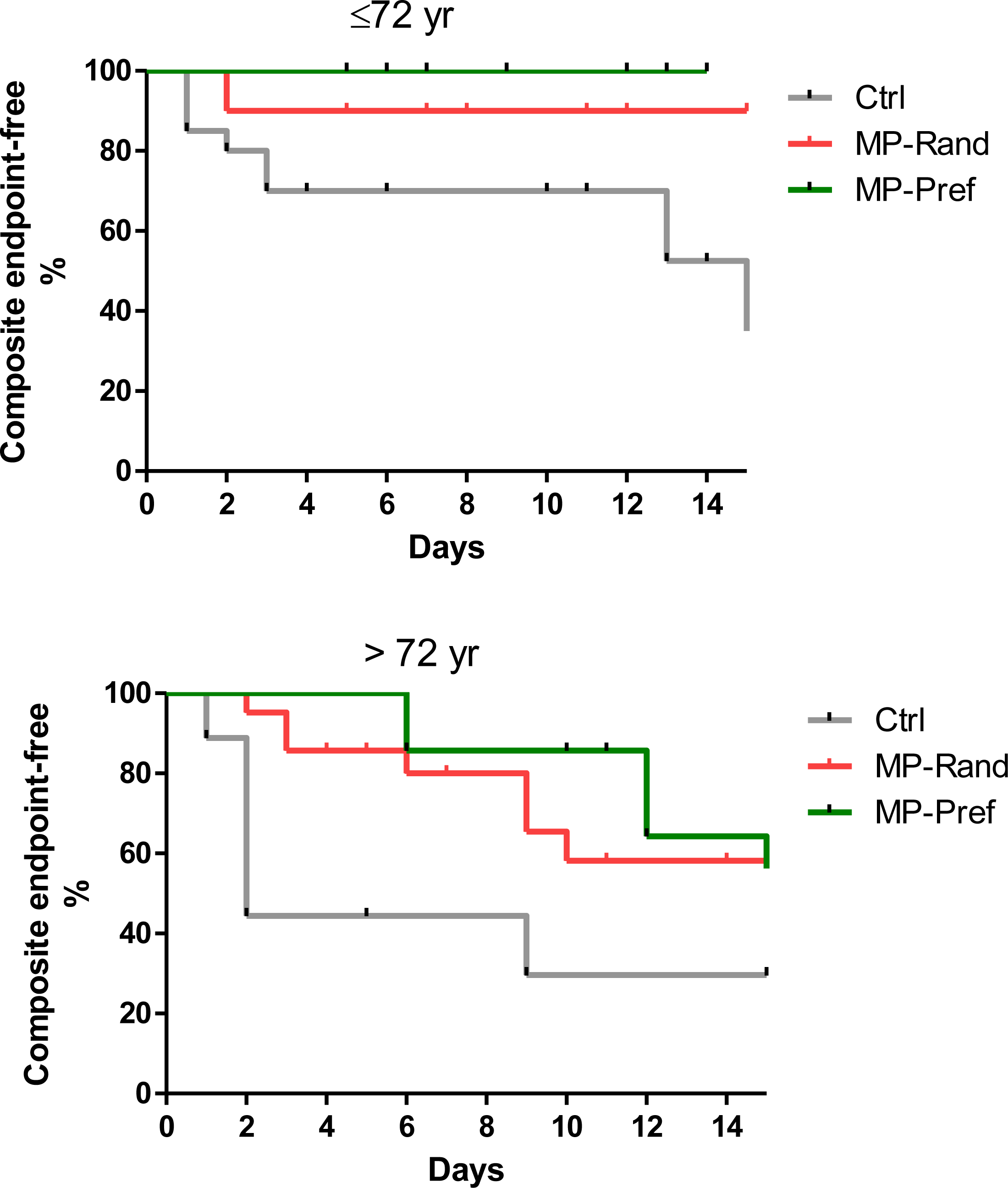
Kaplan-Meier plots showing the probability of not occurring the primary composite endpoint (ICU admission, need of NIV or death) of patients in the control group (grey), randomized MP group (red) and preference MP group (green), stratified by age.

